# Shifting source hubs and the factors associated with cross-country Zika virus transmission: a phylogeographic network analysis, 2012–2025

**DOI:** 10.64898/2026.11.12.26367925

**Authors:** Mingfan Pang, Xinyan Bu, Qiyong Liu, Chao Li, Nimyan Chan, Hanran Ji, Xiaopeng Qi, Yangmu Huang

## Abstract

**Background:** Zika virus local transmission has been reported in at least 97 countries and territories, but the sources, routes, and determinants of cross-country spread remain incompletely understood. We reconstructed a phylogeographic network, characterized source activity across epidemic phases, and examined factors associated with identified cross-country transmission events.

**Methodology/Principal Findings:** We analyzed a time-scaled Nextstrain phylogeny of 1,043 Zika virus genomes collected between February 2013 and March 2026. Parent–child branches assigned to different locations were screened and consolidated into 81 source– destination–year events involving 39 locations. Network analysis characterized temporal and geographic patterns, routes, hubs, and communities. For the analysis of associated factors, 68 events with complete covariate data were compared, using mixed-effects models, with eligible year-matched control records for which no event was identified, with all 68 events retained and a new control set drawn in each of 200 iterations. Asia contributed the earliest event, from Thailand in 2012. Most events occurred within the Americas (79.0%) and were concentrated during 2014–2016, with Brazil as the principal source hub. All six events from 2019 onward originated from Thailand or India and reached destinations in Asia, Europe, and Africa. Thailand’s source activity spanned 2012–2025. Event identification was positively associated with high source-country incidence (odds ratio [OR], 2.66; 95% confidence interval [CI], 1.39–9.69), greater air travel (OR, 2.52; 95% CI, 1.58–5.67), and higher destination-country GNI per capita (OR, 1.70; 95% CI, 1.20–4.50). It was negatively associated with greater geographic distance (OR, 0.11; 95% CI, 0.02–0.20) and higher source-country GNI per capita (OR, 0.64; 95% CI, 0.45–0.94). Fourteen source countries showed elevated residual source propensities, with Brazil and Thailand among the highest.

**Conclusions/Significance:** The transmission network showed a shift in dominant source hubs, from early activity in Asia to the Americas during the 2014–2016 epidemic and back to Asia in recent years. High source-country incidence, greater air travel, and shorter distance can help identify country pairs requiring closer surveillance, alongside established routes in the Americas and persistent or emerging Asian sources, particularly as El Niño may increase vector-borne transmission potential in 2026–2027.

**Author Summary:** Zika virus can spread between countries without being recognized immediately through routine surveillance. We analyzed genetic sequences from 1,043 Zika virus samples and used their evolutionary relationships and geographic information to reconstruct cross-country transmission events. After screening and consolidating records, we identified 81 events involving 39 countries and territories. Source activity changed across epidemic phases. Thailand contributed the earliest event in 2012, while transmission during the 2014–2016 epidemic occurred predominantly within the Americas, with Brazil as the principal source hub. All events identified from 2019 onward originated from Thailand or India, showing the renewed prominence of Asian sources. Cross-country events were more likely to be identified when the source country had high reported Zika incidence, air travel between the countries was greater, and geographic distance was shorter. Several countries remained unusually prominent as sources after the measured factors were taken into account, including the major network hubs Brazil and Thailand. These findings can help health agencies use historical transmission patterns and current epidemiological conditions to identify routes requiring closer surveillance, especially as El Niño may increase the potential for mosquito-borne transmission in 2026–2027.

## 1 Introduction

Zika virus is transmitted primarily by Aedes mosquitoes and can cause congenital Zika syndrome and neurological complications. The 2015–2016 epidemic in the Americas brought Zika to global attention as a major public health threat, and transmission has continued at lower reported levels since the epidemic peak [1, 2]. The 2026 WHO update reported evidence of current or previous autochthonous transmission in 97 countries and territories [3]. Previous studies have examined how Zika virus can spread through environmental suitability, human mobility, and viral evolution. Environmental models have mapped ecological conditions conducive to transmission [4], while models combining air travel and vector distributions have assessed introduction and transmission potential in Africa and the Asia-Pacific region [5]. Genomic studies reconstructed the establishment and cryptic spread of Zika virus in Brazil and the Americas [6], demonstrated that the virus circulated undetected in several American locations for months before the first locally transmitted cases were confirmed [7], and identified multiple introductions into the United States [8]. Earlier and broader phylogeographic studies traced the long-term emergence and movement of Zika virus across Africa, Asia, the Pacific, and the Americas [9, 10].

Existing evidence has largely focused on transmission suitability, importation risk, or epidemic reconstruction in specific settings. What remains less clear is how cross-country viral spread is structured over longer periods, how countries differ in their source and destination roles, and how bilateral and country-level factors are associated with individual transmission events. Climatic anomalies add urgency to this question. Exceptionally warm conditions associated with the 2015 El Niño increased modeled suitability for Zika virus transmission in South America [11]. In September 2026, El Niño was intensifying, with a greater than 90% probability of becoming very strong during the Northern Hemisphere fall and winter of 2026–2027 [12]. Identifying historical routes and their associated factors can therefore provide timely evidence for proactive surveillance.

Accordingly, we aimed to characterize cross-country Zika virus transmission patterns and associated factors to inform risk assessment and early warning in a changing climate. We used a Nextstrain time-scaled phylogeny and ancestral geographic-state annotations to identify phylogenetically inferred cross-country transmission events. Network analysis described when and where these events occurred and how they accumulated into routes, country-level hubs, and communities. The case-control analysis compared observed events with eligible country-pair-year controls to examine source, destination, and bilateral factors associated with event identification.

## 2 Methods

### 2.1 Identification of Phylogenetically Inferred Cross-Country Transmission Events

We conducted a country-level observational study that combined transmission-network analysis with a case-control design. On 17 May 2026, we downloaded a time-scaled phylogeny from the Nextstrain Zika page [13]. The Nextstrain Zika workflow draws primarily on publicly available viral genome sequences and metadata from NCBI GenBank, supplemented by curated sequence data from the United States Virgin Islands. The downloaded tree contained 1,043 viral genomes collected between February 2013 and March 2026.

Internal-node country states were reconstructed in the Nextstrain workflow using maximum-likelihood discrete-trait inference implemented in TreeTime [14, 15]. Using R, we traversed directly connected parent–child branches and identified 131 candidate records of cross-country transmission in which the parent and child nodes were assigned to different countries or territories. When multiple geographic states were available for a node, the country or territory with the highest support was selected. Event year was based on the inferred branch date and could therefore precede the earliest genome sampling date. These candidate geographic transitions represent changes between inferred viral locations and are not counts of notified imported cases or epidemiologically confirmed transmission episodes.

The geographic annotations used by Nextstrain required additional consideration in the identification of cross-country transmission events. The Zika workflow derives geographic metadata primarily from NCBI GenBank and applies standardized geolocation rules and accession-level annotations. For known travel-associated sequences, Nextstrain aims to assign the likely country of infection rather than the country in which the sample was collected. However, when travel history is unavailable or has not been manually curated, a record may retain the sampling location reported in GenBank [13]. To reduce this potential misclassification, we applied geographic eligibility criteria based on WHO documentation of local mosquito-borne transmission through December 2025. Eleven records involving China, Japan, Australia, or Spain were excluded because these locations had imported cases but no documented local mosquito-borne transmission, leaving 120 eligible branches.

To limit the influence of uneven genome sampling and submission, we consolidated records with the same source, destination, and year. Multiple phylogenetic branches representing the same pathway in the same year may reflect the availability of several related genomes rather than independent transmission events. Consolidation removed 39 duplicate records and yielded 81 country-pair-year transmission events for network analysis. Fig 1 shows the selection process and the locations of eligible branches within the phylogeny.

**Fig 1.**
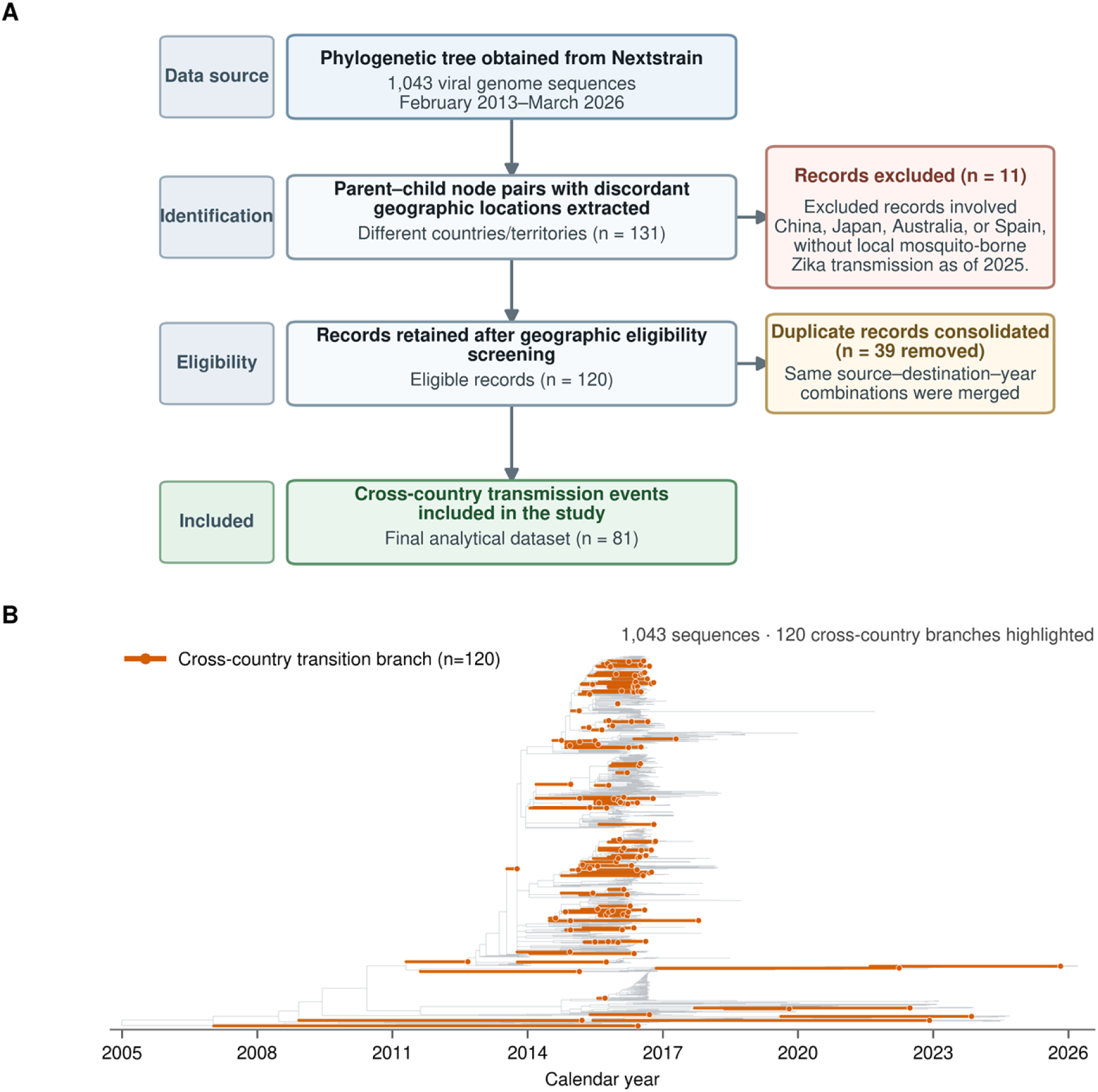
Selection of phylogenetically inferred cross-country Zika virus geographic transitions. (A) Screening of records from a Nextstrain time-scaled phylogeny containing 1,043 genomes sampled from February 2013 to March 2026. Among 131 parent–child branches assigned to different countries or territories, 11 involving China, Japan, Australia, or Spain were excluded to reduce potential misclassification of uncorrected travel-associated cases because these locations had imported cases but no documented local mosquito-borne transmission through 2025. Consolidation of records with the same source–destination–year combination reduced the influence of uneven genome sampling and yielded 81 inferred events from 120 eligible branches. (B) Eligible branches (orange) within the complete phylogeny (grey).

To assess robustness to uncertainty in ancestral geographic-state assignment, we additionally recorded the reconstructed-state probabilities at the parent and child nodes for each cross-country branch. Events were re-consolidated after retaining only branches for which both node probabilities were at least 0.80 or 0.95, and the resulting networks were compared with the primary analysis (S1 Data).

### 2.2 Network Construction and Description

Countries and territories were represented as nodes, and inferred transmission events from source to destination were represented as directed edges. The same directed country pair was aggregated across years into one edge, weighted by the number of its occurrences among the 81 event records. In-degree and out-degree were the numbers of distinct source and destination connections, respectively; total degree was their sum. In-strength and out-strength represented incoming and outgoing event counts, and total strength was their sum. Network density was calculated as the number of observed directed edges divided by all possible directed edges without self-loops. We also calculated reciprocity, clustering coefficient, weakly and strongly connected components, mean shortest path among reachable node pairs, and network diameter.

Source and destination events were summarized annually by continent. Following the United Nations M49 geographic classification, the Americas were further divided into South America, Central America, the Caribbean, and Northern America for the regional transmission matrix [16]. On the geographic network maps, node area was proportional to unweighted total degree, defined as the total number of distinct incoming and outgoing connections, irrespective of the number of events along each connection. Edge width was proportional to the number of country-year transmission events along each directed connection. Community structure was assessed using the Infomap algorithm on the directed network [17] and the Girvan–Newman edge-betweenness algorithm on the undirected projection of the network [18].

### 2.3 Candidate Factors and Analytical Sample

Country-level factors included source-country annual reported Zika incidence, GNI per capita in source and destination countries, the Global Health Security Index (GHSI), and the proportion of land with tropical climate. Annual counts of reported Zika cases were compiled from three source categories: (1) official surveillance and case-reporting data issued by the Pan American Health Organization, national health authorities, and other public health agencies; (2) published literature; and (3) event-based reports from ProMED, WHO Disease Outbreak News, and news media. The case counts were converted to annual reported incidence per 100,000 population, with annual population estimates from the World Bank [19]. GNI per capita, measured using the World Bank Atlas method, was obtained from the World Bank [19]. GHSI is a composite assessment of national capacities to prevent, detect, and respond to epidemic and pandemic threats. We used GHSI scores from the 2019 and 2021 editions, applying the 2019 scores through 2019 and the 2021 scores from 2020 onward [20]. Tropical climate coverage was calculated from the 1-km-resolution Köppen-Geiger raster published by Beck et al. [21] as the proportion of valid land pixels within each country or territory classified as Af (tropical rainforest), Am (tropical monsoon), or Aw (tropical savanna).

Bilateral factors included capital-to-capital geographic distance and annual air travel volume. Distance was represented by the capital-to-capital distance (km) variable from the CEPII Gravity Database [22]. Air travel volume was measured using OAG Traffic Analyser annual passenger-booking counts [23]. Because booking data were available only for 2025, we used 2025 values as a proxy for bilateral mobility throughout the study period. This approach was based on previous findings that international air travel patterns are relatively stable across years, including during the 2015–2016 Zika epidemic, with substantial deviations occurring only during exceptional global disruptions such as the COVID-19 pandemic [24].

The analysis of associated factors was restricted to events with complete covariate data. Thirteen events involving Martinique, Guadeloupe, Saint Barthélemy, French Guiana, the United States Virgin Islands, or American Samoa were excluded because covariate data were unavailable for these locations. The final modeling dataset therefore included 68 observed events involving 31 destination countries and territories. These 68 events were a complete-covariate subset of the 81 events used to construct the network and served as cases in the analysis of associated factors.

### 2.4 Negative-Control Sampling and Model Estimation

The network analysis summarized routes and country-level hubs, whereas the analysis of associated factors compared identified events with eligible controls at the source–destination– year level. The 68 identified events were treated as positive records (cases), whereas negative records (controls) were sampled from eligible source–destination combinations for which no event was identified in the corresponding year. Candidate sources were drawn from the 97 countries and territories listed in the WHO 2026 update [3] as having evidence of current or previous autochthonous Zika virus transmission and were required to have reported at least one Zika case in the corresponding year. Candidate destinations were restricted to the 31 countries and territories represented among the 68 positive events to reduce potential bias arising from geographic differences in genomic surveillance and sequence reporting. Negative-control sampling excluded within-location pairs and any source–destination–year combination already identified as a positive event.

From this candidate pool, year-matched negative records (controls) were sampled at a 1:5 case-to-control ratio. In each iteration, all 68 positive events (cases) were retained, and 340 controls were selected by simple random sampling without replacement. The full candidate pool was restored between iterations, and the sampling procedure was repeated 200 times to assess the stability of estimates across alternative control samples.

We fitted a generalized linear mixed-effects model with a binomial distribution, logit link, and crossed random intercepts for source and destination locations. The random intercepts accounted for residual heterogeneity in each location’s propensity to act as a transmission source or destination after adjustment for the fixed effects, as well as the non-independence of records sharing the same source or destination. The model was specified as:

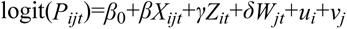

where *P_ijt_* denotes the conditional probability that a cross-country transmission event was identified from source location i to destination location j in year t. The fixed effects comprise bilateral (*X_ijt_*), source (*Z_it_*), and destination (*W_jt_*) covariates; *β*_0_ is the fixed intercept; *β*, *γ*, and *δ* are the corresponding coefficient vectors; and *u_i_*and *v_j_* are random intercepts for source and destination locations, respectively.

To reduce right-skewness, annual air travel volume was transformed as ln(x+1), with 1 added to accommodate records with zero travel volume, while GNI per capita for source and destination locations was natural-log transformed. All continuous covariates were subsequently standardized as Z scores (mean 0 and standard deviation 1) to place them on a common scale. Annual reported Zika incidence in the source location was categorized as low (<5 cases per 100,000 population), medium (5–100 cases per 100,000, including both cutoffs), or high (>100 cases per 100,000), with the low-incidence group as the reference. Odds ratios (ORs) for continuous covariates therefore represent the change in odds associated with a one-standard-deviation increase in the corresponding variable, on the transformed scale where applicable.

Collinearity among candidate covariates was assessed using pairwise Pearson correlation coefficients and variance inflation factors (VIFs). When collinear covariates were identified, variables were selected for inclusion in the final model based on data coverage and epidemiological relevance. For each fixed effect, coefficients were exponentiated to ORs in each of the 200 modeling iterations, and the results were summarized using the median OR and empirical 95% CIs defined by the 2.5th and 97.5th percentiles. Associations were considered statistically significant when the empirical 95% CI excluded the null value (OR = 1). Random intercepts were exponentiated to express each location’s residual effect on the odds relative to a random-intercept value of zero. Analyses were conducted in R version 4.5.1, and generalized linear mixed-effects models were fitted using the lme4 package [25].

## 3 Results

### 3.1 Phylogeographic Network of Cross-Country Zika Virus Spread

#### 3.1.1 Overall Network Structure

We identified 81 phylogenetically inferred cross-country events involving 39 countries and territories. After recurring source–destination connections observed in different years were aggregated, these events represented 65 unique directed routes (Fig 2). Most routes were temporally limited: 51 were observed in only one year, 12 in two years, and two in three years. Brazil-to-United States and Nicaragua-to-Honduras were the only routes observed in three separate years.

**Fig 2.**
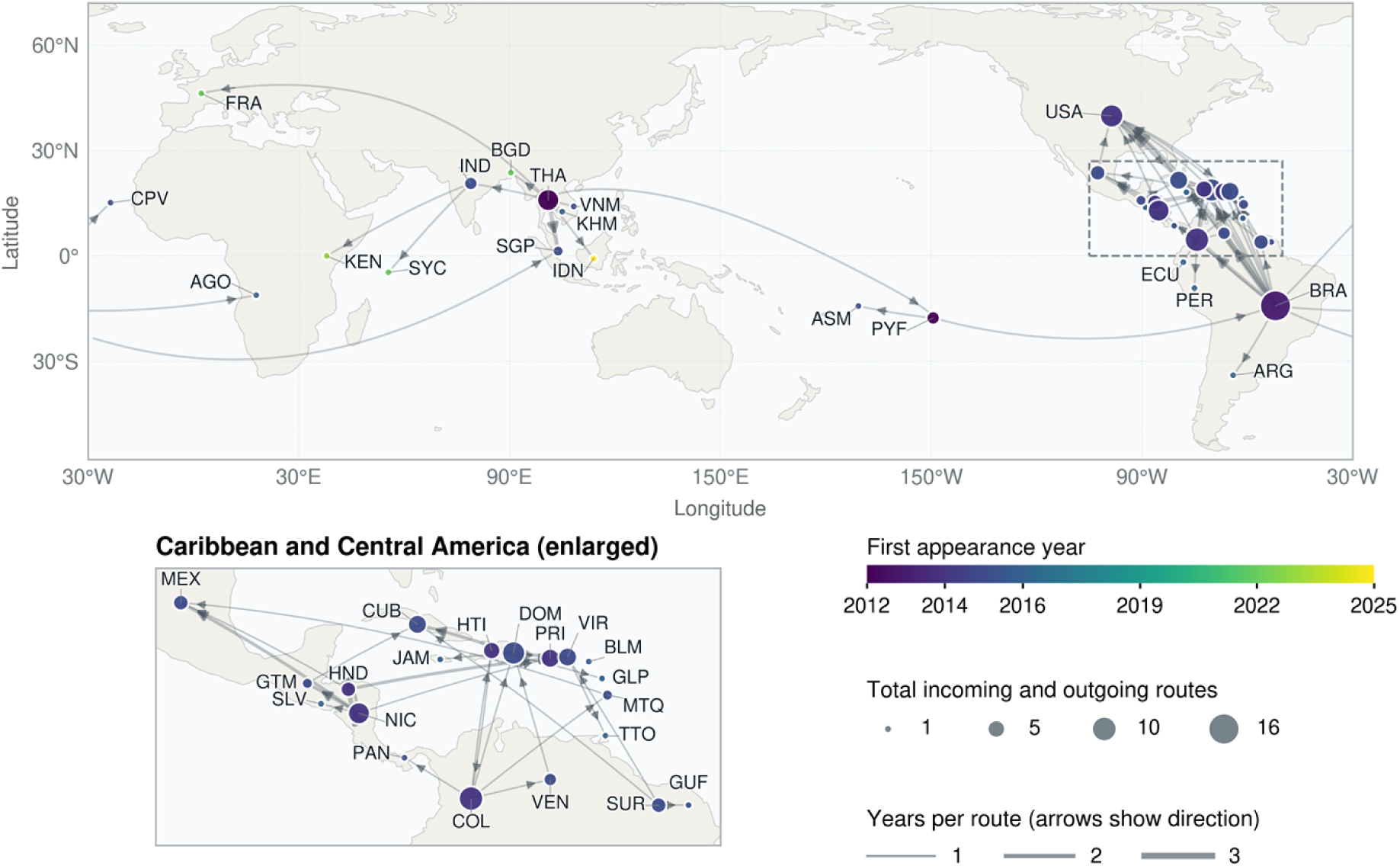
Geographic network of phylogenetically inferred cross-country Zika virus transmission events. The 81 identified cross-country transmission events form 65 unique directed connections among 39 countries and territories. Node area is proportional to unweighted total degree (defined as the total number of distinct incoming and outgoing connections), and node color indicates the first year in which a location appeared in the network. Edge width represents the number of event years for each directed country pair, and arrowheads indicate inferred direction. Labels are ISO 3166-1 alpha-3 codes. ISO code-country/territory correspondence: AGO, Angola; ARG, Argentina; ASM, American Samoa; BGD, Bangladesh; BLM, Saint Barthélemy; BRA, Brazil; COL, Colombia; CPV, Cabo Verde; CUB, Cuba; DOM, Dominican Republic; ECU, Ecuador; FRA, France; GLP, Guadeloupe; GTM, Guatemala; GUF, French Guiana; HND, Honduras; HTI, Haiti; IDN, Indonesia; IND, India; JAM, Jamaica; KEN, Kenya; KHM, Cambodia; MEX, Mexico; MTQ, Martinique; NIC, Nicaragua; PAN, Panama; PER, Peru; PRI, Puerto Rico; PYF, French Polynesia; SGP, Singapore; SLV, El Salvador; SUR, Suriname; SYC, Seychelles; THA, Thailand; TTO, Trinidad and Tobago; USA, United States; VEN, Venezuela; VIR, United States Virgin Islands; and VNM, Vietnam.

Network density was 0.044, meaning that only 4.4% of all possible directed connections among the 39 locations were observed. Despite this global sparsity, the mean shortest-path length among location pairs connected by a directed path was 2.49, indicating that reachable destinations were typically connected to sources within two to three cross-country transmission steps. The global clustering coefficient of the undirected network was 0.263, indicating that 26.3% of connected triplets were closed into triangles, primarily in Central America and the Caribbean. Overall, the network was globally sparse but contained short directed pathways and localized regional clustering.

When edge direction was disregarded, all 39 locations formed a single weakly connected component, indicating that the observed transmission network was globally connected rather than divided into several mutually isolated subnetworks. When direction was retained, however, the network separated into 34 strongly connected components, defined as groups in which every location could reach every other by following directed paths. The largest strongly connected component contained four countries: neighboring Colombia and Venezuela in northern South America, and Haiti and the Dominican Republic, which share the island of Hispaniola in the Caribbean. Network reciprocity was low at 0.092, meaning that only 9.2% of directed connections were matched by a connection in the opposite direction. Reciprocal connections were confined to three pairs: Nicaragua and Honduras, which share a land border; the United States and Cuba, which are separated by the Straits of Florida; and Colombia and Haiti, which lie across the Caribbean Sea. Together, the large number of strongly connected components and the scarcity of reciprocal connections indicate that transmission flows were predominantly one-way.

#### 3.1.2 Temporal and Geographic Distribution

Transmission events showed marked temporal concentration (Fig 3). The earliest events were identified in 2012 and 2013, followed by a rapid increase to 7, 24, and 40 events in 2014, 2015, and 2016, respectively. The 2014–2016 events accounted for 87.7% of the total, and 2016 alone for 49.4%, mainly driven by the epidemic in the Americas. Event frequency declined sharply thereafter, with only two events in 2017. Six events were identified from 2019 onward, spread across 2019–2025 and all originating in Asia, with four from Thailand and two from India. Source activity therefore varied by epidemic phase rather than following a simple unidirectional geographic shift: Asia contributed the earliest source event in 2012, the sharp increase during 2014–2016 was driven primarily by the epidemic in the Americas, and all six events identified from 2019 onward originated from Asia.

**Fig 3.**
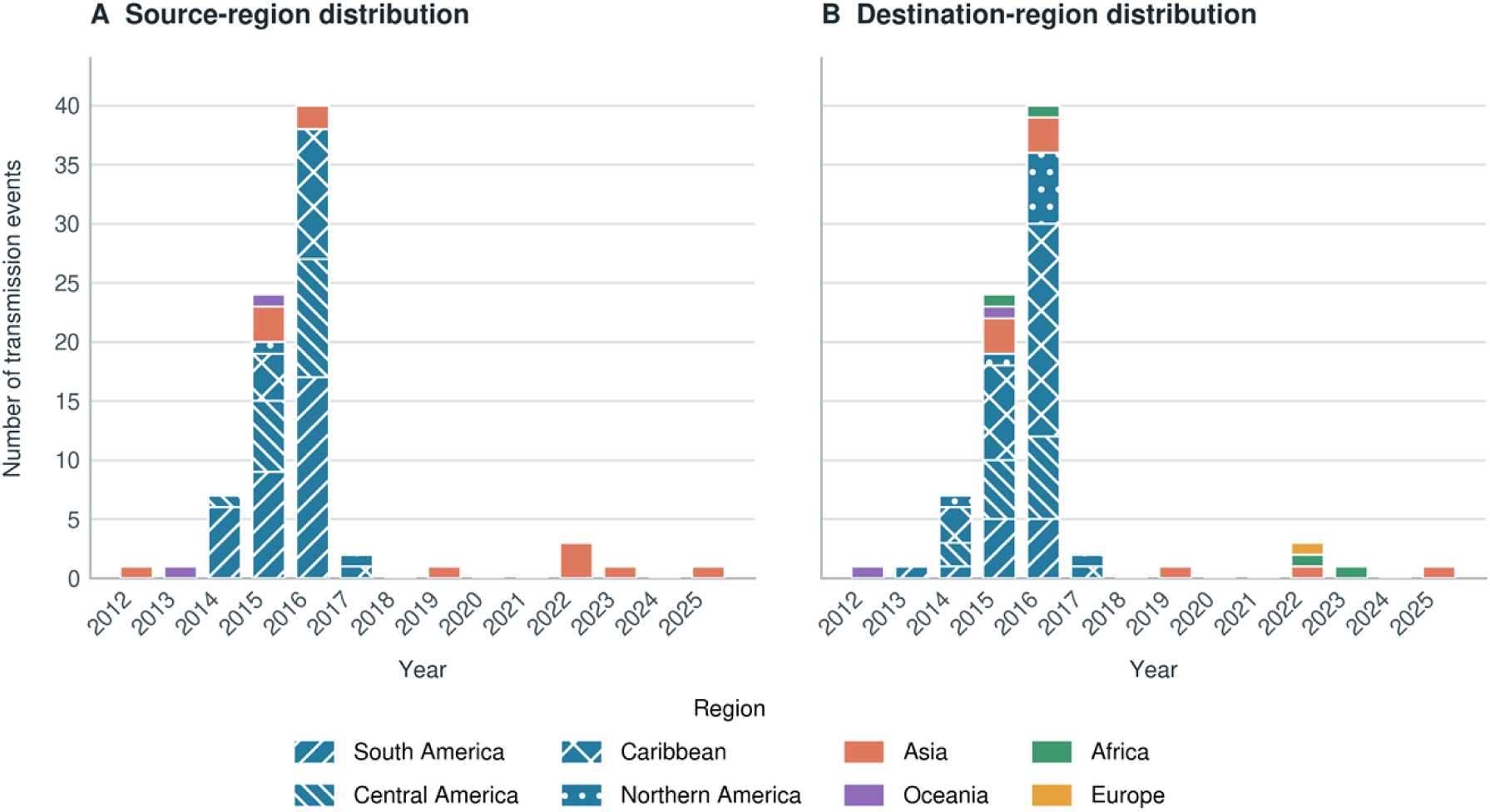
Temporal dynamics of phylogenetically inferred cross-country Zika virus events by continent. Annual source (A) and destination (B) distributions are shown for the 81 inferred events. Bar height represents event count. Colors identify continents; the four American subregions share blue and are distinguished by hatch patterns. Years without identified events are retained. Years refer to inferred event dates.

Geographically, 64 events (79.0%) occurred within the Americas, all during 2014–2017. A further eight events occurred within Asia and one within Oceania. South America was the largest source region, accounting for 32 events, followed by Central America (17), the Caribbean (16), Asia (12), Oceania (2), and Northern America (2). In contrast, the Caribbean was the largest destination region, receiving 30 events, followed by Central America (14), South America (12), Asia (9), Northern America (9), Africa (4), Oceania (2), and Europe (1) (Fig 4). This contrast identified South America as the principal regional origin of transmission and the Caribbean as its main receiving region and hub for onward transmission. Only eight events (9.9%) crossed continents, collectively spanning the pre-epidemic, epidemic, and post-epidemic periods. The earliest followed an eastward sequence, from Thailand to French Polynesia in the South Pacific in 2012 and onward to Brazil in South America in 2013. During the epidemic in the Americas, Brazil was the source of events to Cabo Verde off West Africa in 2015 and to Angola in southern Africa and Singapore in 2016. The most recent intercontinental events were among the post-2019 exports from Asia:

**Fig 4.**
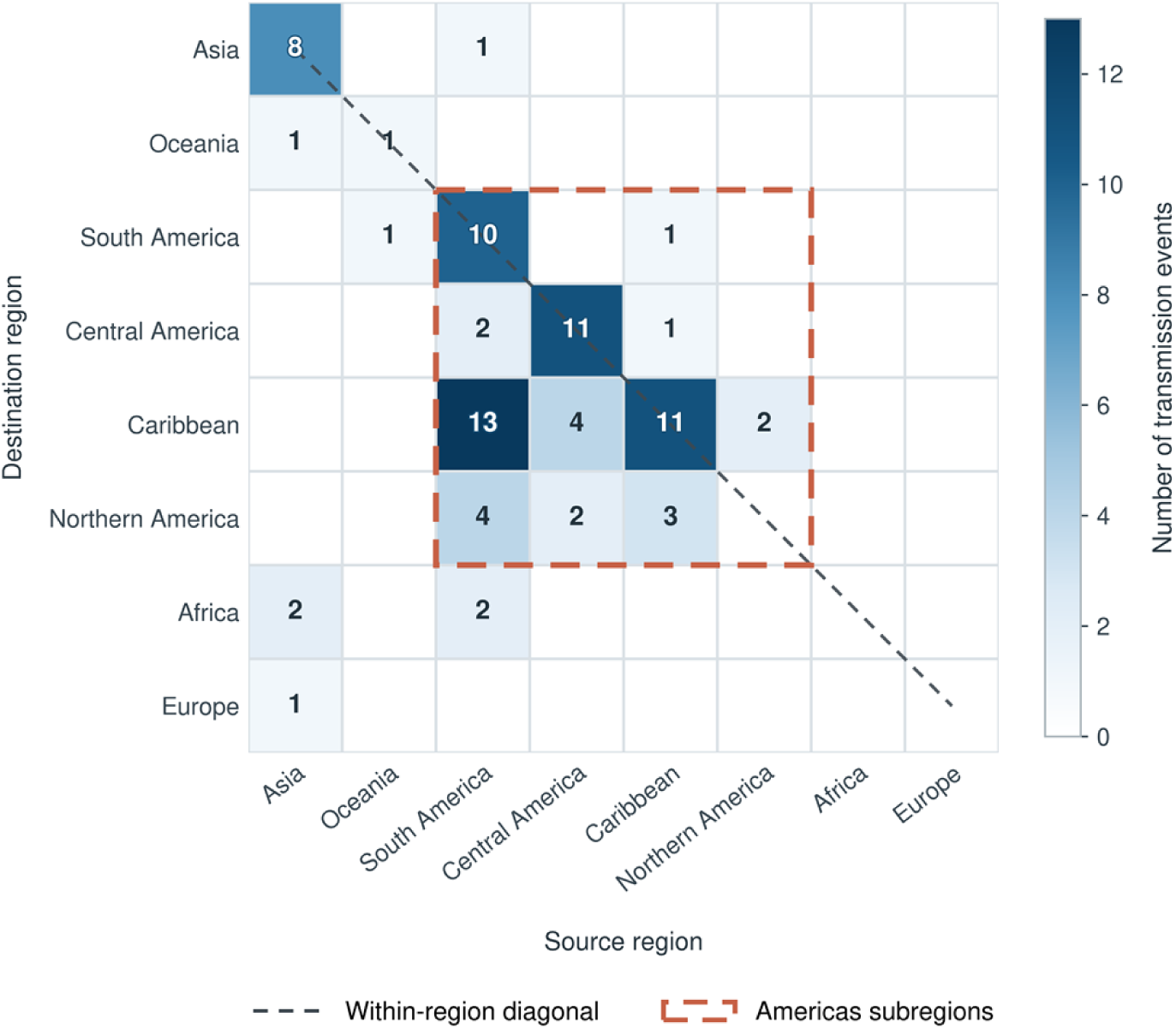
Matrix of phylogenetically inferred cross-regional Zika virus transmission events. Columns indicate source regions and rows indicate destination regions. Cell labels and color intensity represent the number of the 81 inferred events; zero-count cells are white and unlabeled. The dashed diagonal links within-region cells. The dashed orange box identifies South America, Central America, the Caribbean, and Northern America.

Thailand to France and India to Seychelles in the western Indian Ocean in 2022, followed by India to Kenya in East Africa in 2023.

#### 3.1.3 Hubs and Community Structure

Incoming and outgoing patterns revealed three distinct node roles (Table 1). Brazil and Thailand were the two principal source hubs. Brazil had the highest total strength, with 20 outgoing events reaching 15 destinations and only one incoming event, from French Polynesia in 2013. Its outgoing activity was concentrated in 2014–2016. Thailand functioned as an independent Asian source hub, with 10 outgoing events to 8 destinations, six within Asia (India, Singapore, Vietnam, Cambodia, Bangladesh, and Indonesia) and two intercontinental (French Polynesia and France). Notably, its outgoing events occurred intermittently from 2012 to 2025, representing the widest temporal span of outgoing activity among all nodes in the network.

**Table 1.** Countries and territories with the ten highest total strengths].

| **No.** | **Country or territory** | **Total strength** | **In-strength** | **Out-strength** | **In-degree** | **Out-degree** | **Incoming years** | **Outgoing years** |
|---|---|---|---|---|---|---|---|---|
| 1 | Brazil | 21 | 1 | 20 | 1 | 15 | 2013 | 2014, 2015, 2016 |
| 2 | Nicaragua | 13 | 3 | 10 | 2 | 6 | 2014, 2015, 2016 | 2014, 2015, 2016 |
| 3 | Colombia | 11 | 3 | 8 | 2 | 8 | 2014, 2015 | 2014, 2015, 2016 |
| 4 | Dominican Republic | 11 | 3 | 8 | 3 | 6 | 2015, 2016 | 2015, 2016 |
| 5 | United States | 11 | 9 | 2 | 7 | 2 | 2014, 2015, 2016, 2017 | 2015, 2017 |
| 6 | Thailand | 10 | 0 | 10 | 0 | 8 | - | 2012, 2015, 2016, 2019, 2022, 2025 |
| 7 | Honduras | 8 | 3 | 5 | 1 | 3 | 2014, 2015, 2016 | 2015, 2016 |
| 8 | Puerto Rico | 8 | 5 | 3 | 4 | 2 | 2014, 2015, 2016 | 2015, 2016 |
| 9 | Cuba | 7 | 6 | 1 | 5 | 1 | 2015, 2016, 2017 | 2017 |
| 10 | Haiti | 7 | 5 | 2 | 3 | 2 | 2014, 2015, 2016 | 2015, 2016 |

**Note:** Strength is based on the 81 event records; degree is based on distinct directed country pairs. A dash indicates that no corresponding event was observed. The original ordering was retained for ties in total strength.

The United States, Cuba, Haiti, and Puerto Rico were destination-dominant nodes. The United States was the largest destination hub, receiving 9 events from 7 sources, compared with only 2 outgoing events. Cuba, Haiti, and Puerto Rico also received more events than they generated. These nodes represented the principal recipients of transmission from multiple sources.

Nicaragua, Colombia, the Dominican Republic, and Honduras were bidirectional regional hubs. During 2014–2016, they received inputs primarily from Brazil and neighboring locations and subsequently transmitted the virus to destinations across South America, Central America, the Caribbean, and Northern America. All events involving these nodes occurred within the Americas, highlighting their roles as intermediate hubs in regional transmission chains.

Directed Infomap assigned all 39 nodes to a single community, indicating that directed transmission flow did not concentrate into distinct modules. In contrast, edge-betweenness analysis of the undirected network identified six geographically structured communities (Table 2 and Fig 5). Communities 1, 2, 3, and 5 primarily represented transmission within the Americas but differed in their structural roles. Community 1 had a star-like structure centered on Brazil, connecting Brazil with locations in South America, the Caribbean, and Africa. It produced the most outward events among the six communities, highlighting Brazil’s role as the principal source of transmission. Community 2 formed a bridging corridor centered on Colombia and Haiti, linking South America with the Caribbean and Central America through a combination of internal, incoming, and outgoing connections. Community 3 was the largest destination-oriented community, centered on the Caribbean and the United States. It received the greatest number of external inputs and contained the largest number of internal events, indicating substantial transmission both into and within this group. Community 5 was a compact Central American relay community centered on Nicaragua and Honduras, characterized by repeated within-community connections across 2014–2016 and subsequent transmission to the Caribbean and Northern America. Communities 4 and 6 captured the transmission links originating from Asia. Community 4 had a star-like structure centered on Thailand, linking Thailand with destinations in Asia, Oceania, and Europe. Community 6 was the smallest community, comprising three countries linked by two India-originating connections to Africa.

**Fig 5.**
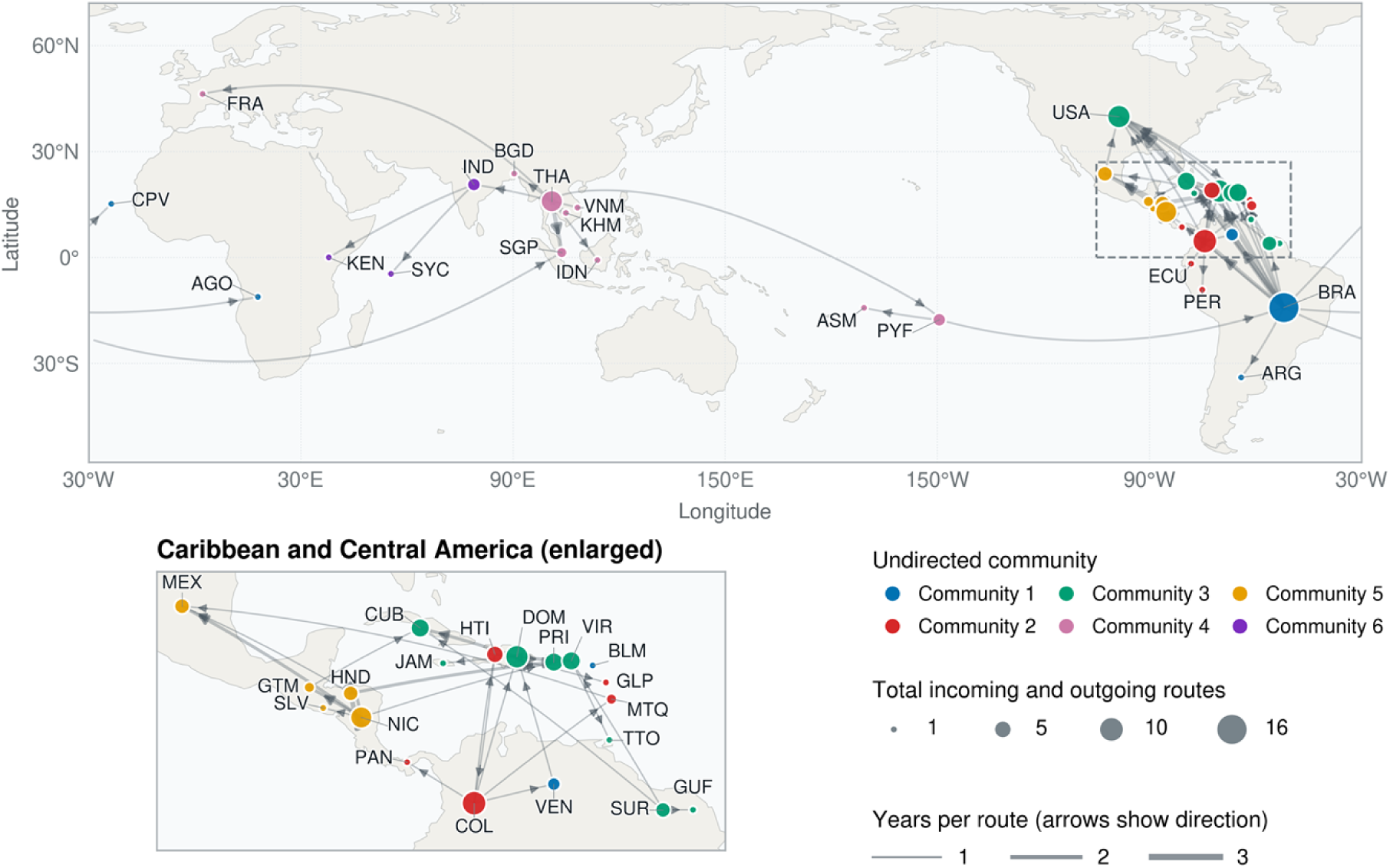
Geographic distribution of communities in the phylogeographic network of cross-country Zika virus spread. Node colors indicate the six communities identified by edge-betweenness analysis of the undirected network; community numbers do not indicate risk levels. Node area represents unweighted total degree. Original directed connections are retained, with edge width proportional to event count and arrowheads indicating inferred direction. The enlarged panel shows the Caribbean and Central America. ISO code definitions are provided in the caption to Fig 2.

**Table 2.** Communities identified by edge-betweenness analysis of the undirected network.

| **Community** | **Countries and territories** | **Within-community events** | **External incoming events** | **External outgoing events** |
|---|---|---|---|---|
| 1 | Brazil, Venezuela, Angola, Argentina, Cabo Verde, Saint Barthélemy | 6 | 2 | 15 |
| 2 | Colombia, Haiti, Martinique, Ecuador, Guadeloupe, Panama, Peru | 7 | 6 | 4 |
| 3 | Cuba, Dominican Republic, Puerto Rico, United States, United States Virgin Islands, Jamaica, French Guiana, Trinidad and Tobago, Suriname | 16 | 17 | 2 |
| 4 | Thailand, French Polynesia, Singapore, Cambodia, Vietnam, Indonesia, Bangladesh, France, American Samoa | 10 | 1 | 2 |
| 5 | Guatemala, Honduras, Mexico, Nicaragua, El Salvador | 11 | 2 | 6 |
| 6 | India, Kenya, Seychelles | 2 | 1 | 0 |

**Note:** A within-community event was defined as an event in which the source and destination belonged to the same community. Incoming and outgoing events were defined from the perspective of each community. Each of the 29 cross-community events was therefore counted once as an outgoing event for the source community and once as an incoming event for the destination community. Community numbers are identifiers and do not indicate risk levels.

### 3.2 Factors Associated With Identified Cross-Country Transmission Events

#### 3.2.1 Variable Selection and Collinearity

Factors associated with event identification were examined at the country-pair-year level using the 68 events with complete covariate data. The Pearson correlation matrix for candidate continuous variables is shown in Fig 6. GHSI and GNI per capita were correlated for both destination (r = 0.72) and source countries (r = 0.56). Given this redundancy and the limited temporal coverage of GHSI, which was available only in the 2019 and 2021 editions, GNI per capita was retained and the GHSI variables were excluded from the model. The final model contained seven factors represented by eight non-intercept fixed-effect coefficients. Across the 200 repeated control samples, the maximum VIF among included variables ranged from 1.202 to 2.408, indicating no evidence of serious collinearity.

**Fig 6.**
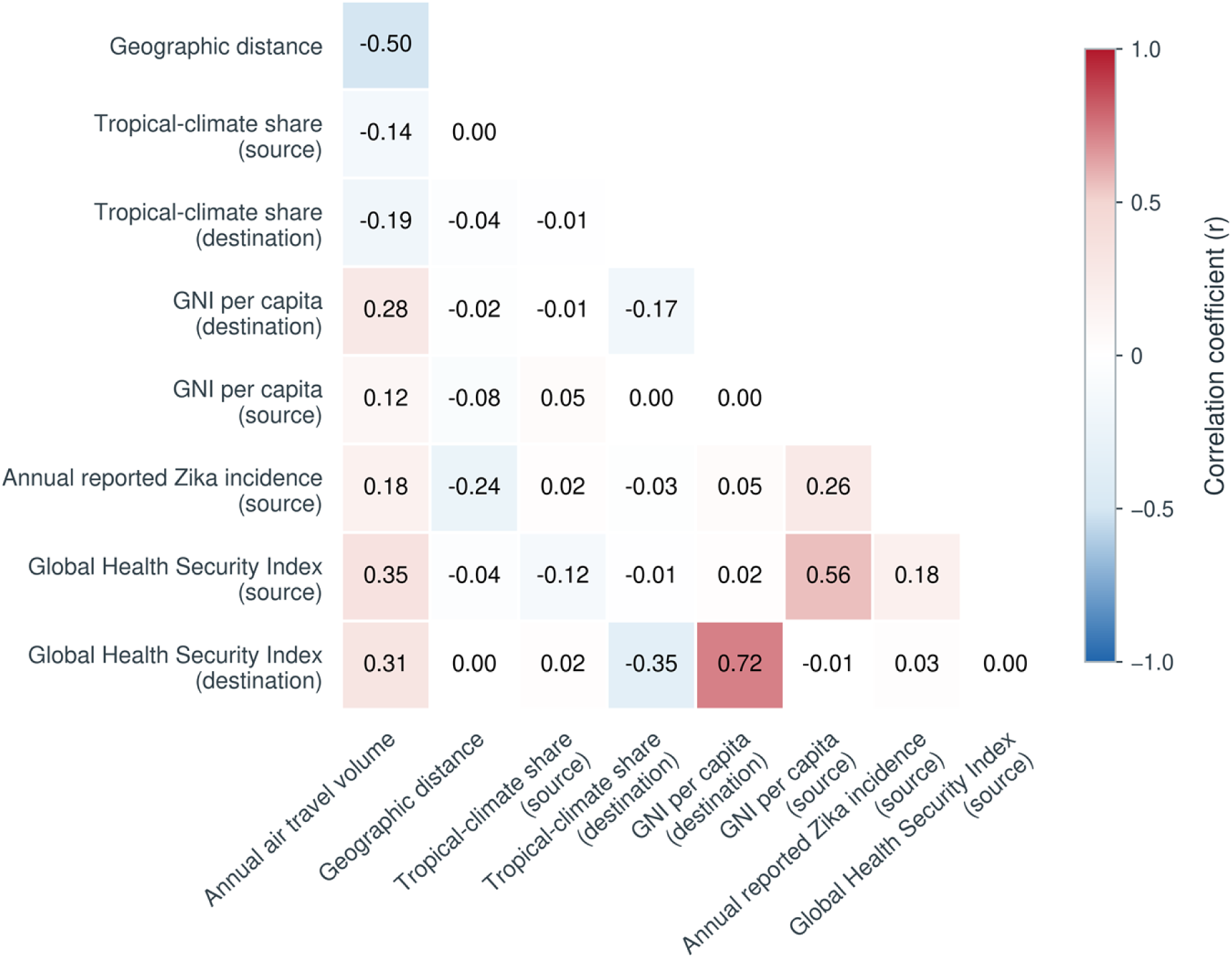
Pearson correlation matrix of candidate continuous variables. Only the lower triangle is displayed; the diagonal and redundant upper triangle are omitted. Red indicates positive and blue negative correlations, with intensity proportional to the absolute coefficient. Numbers are Pearson correlation coefficients.

#### 3.2.2 Fixed Effects

Fixed-effect results are shown in Fig 7, with complete estimates in S1 Table. High versus low source-country incidence (>100 versus <5 cases per 100,000) was positively associated with event identification (OR, 2.66; empirical 95% CI, 1.39–9.69). Higher odds were also associated with annual air travel volume (OR, 2.52; 95% CI, 1.58–5.67) and destination-country GNI per capita (OR, 1.70; 95% CI, 1.20–4.50). In contrast, geographic distance (OR, 0.11; 95% CI, 0.02–0.20) and source-country GNI per capita (OR, 0.64; 95% CI, 0.45–0.94) were associated with lower odds of an identified event. The remaining fixed effects showed no clear association, with confidence intervals spanning the null: medium versus low source-country incidence (OR, 0.62; 95% CI, 0.16–1.52) and the proportions of tropical climate in source (OR, 1.01; 95% CI, 0.75–1.40) and destination countries (OR, 1.26; 95% CI, 0.90–1.83).

**Fig 7.**
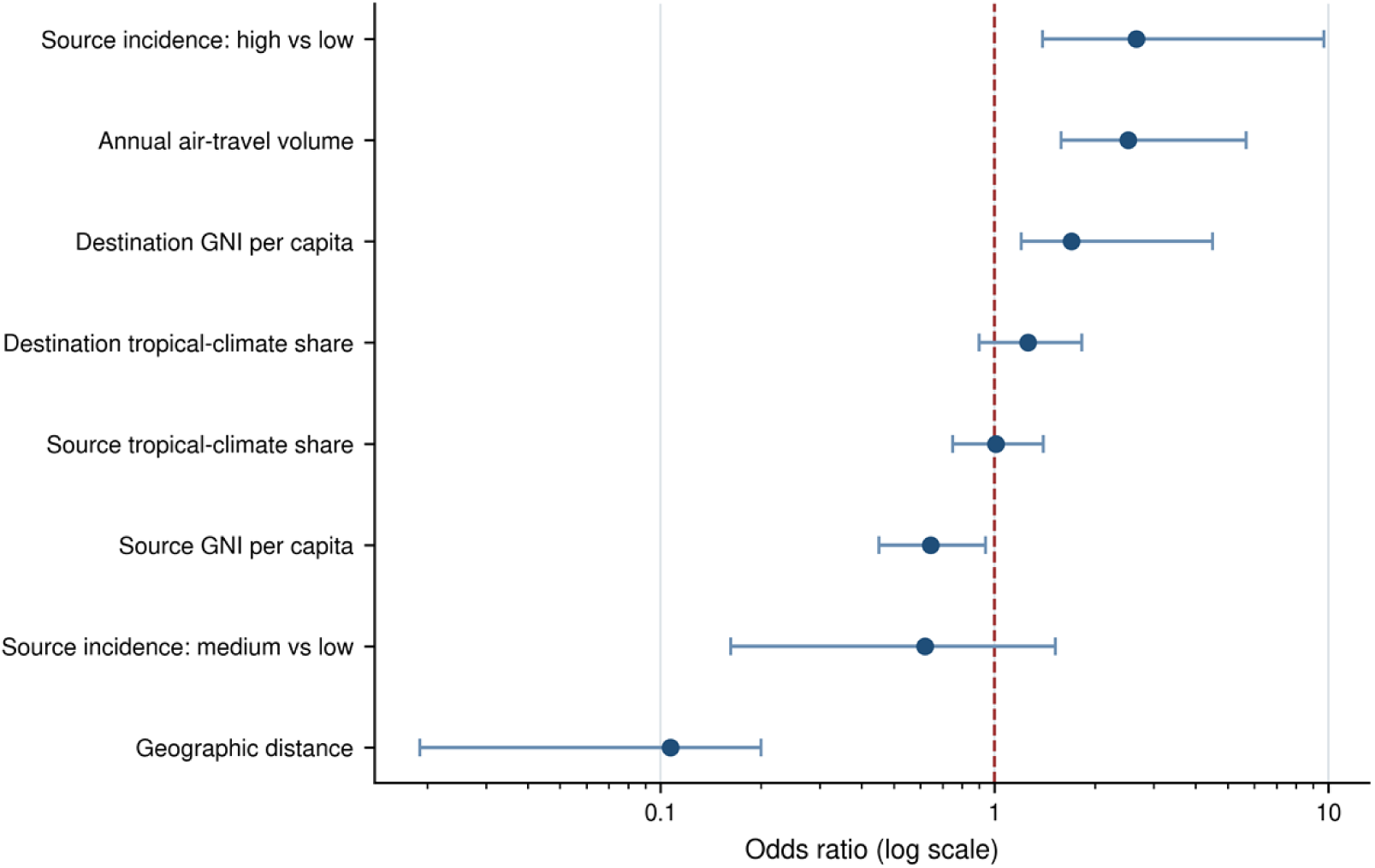
Fixed-effect estimates of factors associated with identified cross-country Zika virus transmission events. Points and horizontal lines represent median odds ratios (ORs) and empirical 95% confidence intervals across 200 repeated control samples. Source-country annual reported incidence was categorized as low (<5 cases per 100,000), medium (5–100), or high (>100), with low as the reference category. Continuous-predictor ORs represent a one-standard-deviation increase after log transformation and standardization. The vertical dashed line marks OR = 1; the horizontal axis is on a logarithmic scale.

#### 3.2.3 Source- and Destination-Country Random Effects

After adjustment for the fixed effects, exponentiated source-country random-intercept estimates (S2 Table) exceeded 1, with empirical 95% CIs excluding 1, for 14 of the 75 source countries, and were ranked highest for Brazil, Nicaragua, Thailand, the Dominican Republic, and India. Brazil and Thailand, the two principal source hubs identified in the network, ranked first and third, respectively, in residual source propensity. This concordance connects their node-level network roles with the event-level mixed model while indicating that the measured covariates did not fully account for their prominence. Given the small number of positive events and the wide, mutually overlapping country-level intervals, these estimates should be compared by relative ranking rather than by the magnitude of individual point estimates. By contrast, the empirical 95% CIs for all destination countries contained 1, indicating no clear residual destination propensity.

## 4 Discussion

This study reconstructed more than a decade of phylogenetically inferred cross-country Zika virus spread and examined factors associated with identification of individual transmission events. Three findings stand out. First, source activity varied across epidemic phases. Asia contributed the earliest inferred event through Thailand in 2012; Brazil and the wider Americas dominated the high-volume 2014–2016 epidemic; and all events identified from 2019 onward originated from Thailand or India in Asia. Second, the network was highly concentrated: it was sparse and predominantly directional, most routes appeared in only one year, and a small number of source hubs and recurrent routes carried much of the activity. Third, at the event level, transmission events were more likely to be identified when source-country incidence was high, bilateral air travel was substantial, and geographic distance was short; source-country random effects were also concordant with the principal hubs identified in the network.

### 4.1 Phase-Specific Source Patterns and Shifting Hubs

Cross-country Zika virus transmission events were concentrated in the Americas during 2014–2016, with South America serving as the main source region and the Caribbean receiving the largest number of incoming events. This pattern is consistent with genomic evidence on the establishment and rapid spread of Zika virus in the Americas [6]. Bayesian phylogeographic analyses further reconstructed the underlying directed movements: after its establishment in northeast Brazil, the virus reached Haiti around mid-2014 and subsequently Colombia, the Dominican Republic, and other American countries [33, 34]. Genomic analyses have also shown that the virus circulated undetected in several locations before locally transmitted cases were first reported [7], underscoring how case reports may lag behind actual transmission. By applying a consistent country-pair-year unit, our analysis describes a historical network in which intensive within-Americas transmission was connected to a smaller number of intercontinental routes. This event-based network approach follows frameworks developed for other viruses, in which global influenza spread was represented as a network of seeding events between regions [31] and viral genetic data were combined with human transportation data to explain dissemination [32].

Brazil was the leading source node, the United States was predominantly a destination node, and several Caribbean countries and territories played both incoming and outgoing roles. Genomic evidence of multiple introductions into the United States [8] supports the observed multi-source pattern. The dense Caribbean connections in our network further indicate that the region served not only as a destination but also as an intermediate hub for onward transmission.

Asia was not a newly emerging source after 2019. Thailand generated the earliest inferred source event in the network in 2012, followed by the inferred French Polynesia-to-Brazil event in 2013. This sequence is consistent with the broad movement of the Asian lineage through the Pacific to the Americas described in phylogeographic research [9, 10], although unsampled locations or transmission steps may lie between the inferred events. The post-2019 pattern instead represents a renewed concentration of identified source activity in Asia after the epidemic peak in the Americas. All six events identified from 2019 onward originated from Thailand or India, linking Asia to Europe and Africa as well as destinations within Asia. India’s emergence as a recent source is consistent with documented outbreaks since 2018 caused by Asian-lineage viruses and with evidence of continued circulation across multiple Indian states [26, 27]. Independent surveillance evidence supports Thailand’s persistent source role: nationwide testing in 2016–2017 confirmed widespread focal transmission across 29 of 76 provinces, and time-resolved phylogenetic analysis indicated that Zika virus had circulated in Thailand at a low but sustained level since at least 2002, with viral diversity approaching endemic patterns [28]. A comprehensive phylogenomic analysis likewise identified Thailand as a crucial hub for Zika virus spread across Asia [36].

### 4.2 Factors Associated With Identified Cross-Country Transmission Events

The analysis of associated factors complements the network analysis by examining the conditions associated with the individual transmission events that collectively produced the observed routes and hubs. Annual air travel volume was positively associated with event identification, whereas geographic distance was negatively associated, highlighting the joint roles of human mobility and spatial proximity. These findings provide empirical support for early travel-based risk models, which anticipated that international air movement could introduce Zika virus into vector-suitable areas [5].

High source-country incidence was significantly associated with an identified cross-country transmission event, whereas medium incidence did not differ significantly from low incidence. This pattern suggests a possible threshold effect: increases within the low-to-medium range may not markedly change exportation risk, but once incidence exceeds a higher threshold, the number of infections available for export, and thus the probability that a cross-country transmission event is identified, may rise substantially. These categories do not define a precise biological threshold, but they suggest that rapid escalation into the high-incidence range is a useful signal for surveillance in destination countries.

Two mechanisms could underlie such a threshold pattern. First, the pool of viremic travelers grows approximately with the product of source-country incidence and travel volume, so the probability that at least one infection is exported may follow a saturating function of pool size, remaining low while the pool is small and then rising steeply once incidence enters the high range. Second, reported incidence partly reflects reporting propensity: countries experiencing recognized outbreaks intensify case finding and sequencing, which raises both recorded incidence and the likelihood that an exported infection is detected and genetically linked to its source. These mechanisms cannot be separated with the present design, but they yield a testable prediction: if the threshold reflects a true biological process, exportation probability should rise steeply within a defined incidence range once continuous case counts and dense traveler sampling become available; if it is purely an artifact of reporting propensity, the pattern should attenuate once reporting effort is held constant.

Tropical climate coverage in source and destination countries was not significantly associated with transmission. This null result may reflect the study design. Both candidate pools were restricted: source countries were required to have current or previous local transmission, and destination countries were limited to those represented among the positive events. Broad ecological suitability was therefore already common across the sampled countries, and tropical climate coverage varied little within the analysis sample. Moreover, a static measure cannot capture year-specific variation in climatic suitability. This finding therefore does not imply that climate is unimportant, but rather that a time-invariant national measure with limited within-sample variation is insufficient for detecting climatic effects. A previous study projected that longer-term climate change would redistribute the geographic and seasonal risk of Aedes-borne viruses [29], and with a very strong El Niño forecast for 2026–2027 [12], near-term climatic influences on Zika virus transmission warrant close attention.

Source-country GNI per capita was negatively associated with transmission events, whereas destination-country GNI per capita was positively associated. At the source, lower national income is often accompanied by poorer housing, greater vector exposure, and less intensive vector control, which can increase local transmission intensity and thus the pool of infections available for export. At the destination, higher income reflects not only stronger international travel connections but also greater diagnostic and genomic surveillance capacity, so events involving wealthier destinations may be both more likely to occur and more likely to be detected. The source-side association is consistent with an ecological assessment in which lower regional GDP per capita predicted greater Zika virus transmission potential [35].

Because a source node’s strength reflects the accumulation of individual country-pair-year events, the fixed-effect associations identify general conditions through which countries may acquire multiple outgoing routes. After adjustment for these factors, 14 source countries retained residual source propensities whose empirical 95% CIs exceeded 1, ranked highest for Brazil, Nicaragua, Thailand, the Dominican Republic, and India; no destination-country interval excluded 1. Brazil and Thailand nevertheless remained first and third in the source ranking, indicating that their hub roles were only partly captured by incidence, air travel, distance, income, and climate. Their remaining prominence may reflect epidemic timing, subnational vector density, sustained local circulation, air-travel patterns not captured by the 2025 proxy, non-air movement, surveillance and sequencing capacity, and other unmeasured factors.

Taken together, the historical network and factor analysis can inform risk-based surveillance prioritization at three levels. At the source level, surveillance can track rapid increases in incidence and in residual source propensity among major sources. At the route level, importation monitoring can prioritize country pairs with substantial travel volume or historically recurrent transmission. At the destination level, travel-associated case detection can be strengthened on these routes. During climatic anomalies that increase vector suitability, such prioritization can direct vector monitoring, genomic sequencing, and rapid cross-country data sharing toward settings where diagnostic and sequencing capacity is limited.

### 4.3 Study Contributions and Limitations

This study integrates phylogenetic evidence, network analysis, and a case-control mixed model within a single framework. The two analytical components address complementary levels: network metrics aggregate identified transmission events to characterize routes and hubs, whereas the mixed model estimates factors associated with identification of those events and captures residual source and destination propensity. Together, they connect historical reconstruction with prospective risk alerts and help identify surveillance priorities.

The study has several limitations. The analysis was conditioned on a single published tree, and uncertainty in the tree topology itself was not propagated. Ancestral geographic-state assignments vary in confidence across the tree. To assess robustness to this uncertainty, we re-consolidated events from branches whose parent- and child-node state probabilities were both at least 0.80 or 0.95 (S1 Data): 60 of 81 events (74.1%) were retained at 0.80 and 53 (65.4%) at 0.95. Events with lower assignment confidence were concentrated among deep nodes of the 2014–2016 expansion in the Americas, whereas all six events from 2019 onward were retained at the 0.80 threshold and four of the six at 0.95. The eight leading source countries and the phase-specific pattern—dominance of the Americas during 2014–2016 and Asian origin of all identified post-2019 events—were unchanged at both thresholds, indicating that the main conclusions are robust to geographic-assignment uncertainty.

Public genome coverage varied across countries and years, and the analysis was limited to samples collected and made publicly available from February 2013 onward, which may have affected event detection and network structure [30]. Terminal-node country annotations may represent either a curated infection location or, when travel history was unavailable or had not been manually reviewed, the sampling location reported in GenBank. Excluding records involving countries without documented local mosquito-borne transmission reduced the likelihood that uncorrected imported cases were interpreted as transmission locations, although it may not have eliminated all such misclassification. Coverage and reported incidence alike were shaped by surveillance and sequencing intensity, which were uneven across countries and declined after the 2015–2016 epidemic [2]. Even at the epidemic peak, the majority of infections in the Americas were not reported, with estimated reporting probabilities varying substantially by year [37, 38]. The concentration of identified events in Asia after 2019 may consequently reflect, in part, where surveillance and sequencing remained most active rather than the complete absence of transmission elsewhere. Thailand’s prominence is consistent with its comparatively strong arbovirus surveillance and genomic capacity [28], which makes it both a genuine source and a sentinel through which wider circulation across Southeast Asia becomes visible. The events should therefore be interpreted as phylogenetically inferred geographic transitions rather than as evidence that transmission did not occur between other country pairs, and observed events may condense longer chains whose intermediate stops were never sampled.

Finally, the factor analysis has its own limitations. Negative-control sampling was restricted to the candidate pools, so any residual bias from unmeasured factors influencing pool membership would be toward the null. Covariate measurement adds two further constraints: 2025 air travel data served as a proxy for mobility during the study period, and the climate covariates, defined as fixed climate-type proportions, did not vary over the study period, so climatic effects could be identified only from cross-country contrasts. Future studies should expand genome coverage and incorporate annual mobility, time-varying climate variables, and vector-density measurements to evaluate the main associations and improve the precision of risk alerts.

## 5 Conclusions

Source activity varied markedly across the study period. Asia contributed the earliest inferred event through Thailand in 2012; intensive within-Americas activity dominated the 2014–2016 epidemic, with Brazil as the principal source; and all events identified from 2019 onward originated from Thailand or India in Asia. The events forming the network were concentrated along a small number of routes and were more likely to be identified when source-country incidence was high, bilateral air travel was substantial, and geographic distance was short. Fourteen source countries retained elevated residual source propensities after adjustment, including the principal network hubs Brazil and Thailand, indicating that the measured factors partly, but not fully, accounted for their prominence. Surveillance should continue monitoring established routes in the Americas while prioritizing persistent and emerging Asian sources, issuing alerts when source-country incidence enters the high-incidence range, and strengthening case detection and genomic data sharing across these routes, particularly in resource-constrained settings.

## Data Availability

The time-scaled phylogeny and associated metadata analyzed in this study are publicly available from Nextstrain (https://nextstrain.org/zika) [13], and the corresponding ingest and phylogenetic workflow code is available at https://github.com/nextstrain/zika. The GenBank accessions of the 1,043 genomes in the analyzed tree version are listed in S3 Data. The 81 reconstructed source–destination–year events, with supporting branch counts, geographic-assignment confidence, and sensitivity-retention indicators, are provided in S1 Data, and the underlying 120 branch-level records in S4 Data. The case-control analysis dataset (S2 Data) contains the 68 positive events, the full candidate pool of eligible negative-control records, and all covariate values. Variable definitions and data sources are documented in S1 Text. Air passenger-booking data were obtained from OAG Traffic Analyser [23] and are proprietary; the derived annual bilateral passenger counts used in the analysis are included in S2 Data. All other data sources are publicly available as cited [3, 16, 19–22].

## Data Availability

The time-scaled phylogeny and associated metadata analyzed in this study are publicly available from Nextstrain (https://nextstrain.org/zika) [13], and the corresponding ingest and phylogenetic workflow code is available at https://github.com/nextstrain/zika. The GenBank accessions of the 1,043 genomes in the analyzed tree version are listed in S3 Data. The 81 reconstructed source–;destination–year events, with supporting branch counts, geographic-assignment confidence, and sensitivity-retention indicators, are provided in S1 Data, and the underlying 120 branch-level records in S4 Data. The case-control analysis dataset (S2 Data) contains the 68 positive events, the full candidate pool of eligible negative-control records, and all covariate values. Variable definitions and data sources are documented in S1 Text. Air passenger-booking data were obtained from OAG Traffic Analyser [23] and are proprietary the derived annual bilateral passenger counts used in the analysis are included in S2 Data. All other data sources are publicly available as cited [3, 16, 19–;22].

## Supporting information

**S1 Table. Fixed-effect estimates from repeated mixed-effects logistic regression models of cross-country Zika virus transmission events**

**S2 Table. Source countries with elevated residual transmission-source propensity in the repeated mixed-effects logistic regression models.**

**S1 Data. Reconstructed source-destination-year cross-country Zika virus transmission events (n = 81), with supporting branch counts, geographic-assignment confidence, and sensitivity-retention indicators. (CSV)**

**S2 Data. Case-control analysis dataset: the 68 positive events and the 6,642 eligible negative-control candidate records, with all covariate values. (CSV)**

**S3 Data. GenBank accessions of the 1,043 Zika virus genomes in the analyzed Nextstrain tree version. (CSV)**

**S4 Data. Branch-level cross-country transmission records (n = 120) underlying the consolidated events in S1 Data. (CSV)**

**S1 Text. Data dictionary for S1 Data, S2 Data, S3 Data, and S4 Data.**

